# Ability of Bifidobacterium breve 702258 to transfer from mother to infant: the MicrobeMom randomised controlled trial

**DOI:** 10.1101/2023.03.28.23287708

**Authors:** Rebecca L. Moore, Conor Feehily, Sarah Louise Killeen, Cara A. Yelverton, Aisling A. Geraghty, Calum J Walsh, Ian J. O’Neill, Ida Bush Nielsan, Elaine M. Lawton, Rocio Sanchez Gallardo, Sai Ravi Chandra Nori, Fergus Shanahan, Eileen F. Murphy, Douwe Van Sinderen, Paul D. Cotter, Fionnuala M. McAuliffe

## Abstract

**Background:** The composition of the infant microbiome can have a variety of short- and long-term implications for health. It is unclear if maternal probiotic supplementation in pregnancy can impact infant gut microbiome.

**Objective:** The aim of our study was to investigate if maternal supplementation of a formulation of Bifidobacterium breve 702258 from early pregnancy until three months postpartum could transfer to the infant gut.

**Study design:** This was a double-blinded placebo controlled randomised-controlled trial of B. breve 702258 (minimum 1×10^9^ colony forming units) or placebo taken orally from 16-weeks’ gestation until three-months postpartum in healthy pregnant women. The primary outcome was presence of the supplemented strain in infant stool up to 3 months of life, detected by at a least two of three methods, i.e., strain specific PCR, shotgun metagenomic sequencing, or genome sequencing of cultured B. breve. 120 individual infants’ stool samples were required for 80% power to detect a difference in strain transfer between groups. Rates of detection were compared using Fishers exact test.

**Results:** 160 pregnant women with average age 33.6 (3.9) years, mean BMI of 24.3 (22.5, 26.5) kg/m^2^ and 43% with nulliparity (n=58) were recruited from September 2016 to July 2019. Neonatal stool samples were obtained from 135 infants (65 in intervention and 70 in control). The presence of the supplemented strain was detected through at least two methods (PCR and culture) in two infants in the intervention group (n=2/65, 3.1%) and none in the control group (n=0, 0%), *p* = 0.230.

**Conclusion:** Direct strain transfer from mothers to infants of B. breve 772058 occurred, albeit infrequently. This study highlights potential for maternal supplementation to introduce microbial strains into the infant microbiome.

**Trial registration number:** ISRCTN53023014

## Introduction

The composition of the infant microbiome is important for long-term health^1^. During and after birth, the infant gut is colonised through a variety of means^2^. Members of the genus *Bifidobacterium* are among the earliest and found in lower levels in infants born via caesarean versus vaginal delivery^3–5^.In addition, breast feeding influences the abundance of *Bifidobacterium* in infant stool^6^. Due to their ‘Generally Regarded as Safe’ status, the commercial potential of certain *Bifidobacterium* spp. has driven much of the investigation into their purported benefits, which include positive effects on diarrhoeal disease^7, 8^, resistance to microbial infection^9, 10^, risk of colorectal cancer^11, 12^, and the immune system^13, 14^. Commercially available strains report effects from reducing inflammation^15, 16^ and intestinal permeability in rats^17^, to producing beneficial compounds such as gamma-aminobutyric acid^18^, antimicrobial peptides^19^ and conjugated linoleic acid (CLA)^20–22^.

It is unclear clear if probiotic supplementation to the mother during pregnancy can impact on the health of her infant^2, 4, 23^. Previous studies included short durations of maternal antenatal probiotic supplementation and limited antenatal investigation of potential factors including mother-to-infant transfer^23^. The aim of our study was to investigate whether maternal supplementation of *B. breve* 702258 from early pregnancy until 3-months postpartum, would result in transfer of the strain to the infant gut. This bifidobacterial strain was chosen as it has probiotic potential, given it has been shown to influence fatty acid composition by increasing CLAs. This has the potential to positively influence infant health, given CLAs may influence neurodevelopment and inflammation^24–26^.

## Materials and methods

### Study overview

The MicrobeMom study was a single-centre, double-blind, placebo-controlled, randomised controlled trial (RCT) of a supplement with *B. breve* 702258, a strain previously shown to have lipid-modulating potential, in healthy pregnant participants. The trial was prospectively registered with the ISRTCN in August 2016 (ISRCTN53023014, https://doi.org/10.1186/ISRCTN53023014). The study is reported according to the EQUATOR network using the CONSORT guidelines (supplemental file 1)^27^. The study did not include patient or public involvement in trial design.

### Ethical approval

The study received institutional ethical approval from the National Maternity Hospital in February 2016 (EC 35.2015). Written informed maternal consent was obtained and study processes were completed in accordance with the Declaration of Helsinki^28^. As this intervention was a nutritional supplement and not a medication, neither approval from Health Products Regulatory Authority, nor a data monitoring committee was mandated. Study recruitment commenced on 15^th^ of September 2016 and the last visit was completed on 12^th^ July 2019.

### Participant selection

Pregnant individuals were screened for eligibility for the study at their first antenatal visit (between ten- and 15-weeks’ gestation), through review of their medical chart. Individuals were eligible if they had a body mass index (BMI) between 18·5 and 35 kg/m^2^, were aged over 18 years, capable of giving informed consent, and had an adequate level of English language to enable study comprehension. Exclusion criteria included history of gestational diabetes (GDM), diabetes mellitus or pre-diabetes, multiple pregnancy, fetal anomaly, previous perinatal death, any medical condition requiring treatment or unwillingness to limit intake of other probiotic food or supplements during the trial (Fig 1). Out of n=3,548 women assessed for eligibility, n=160 were randomised to the intervention (Fig. 1).

**Figure 1:**
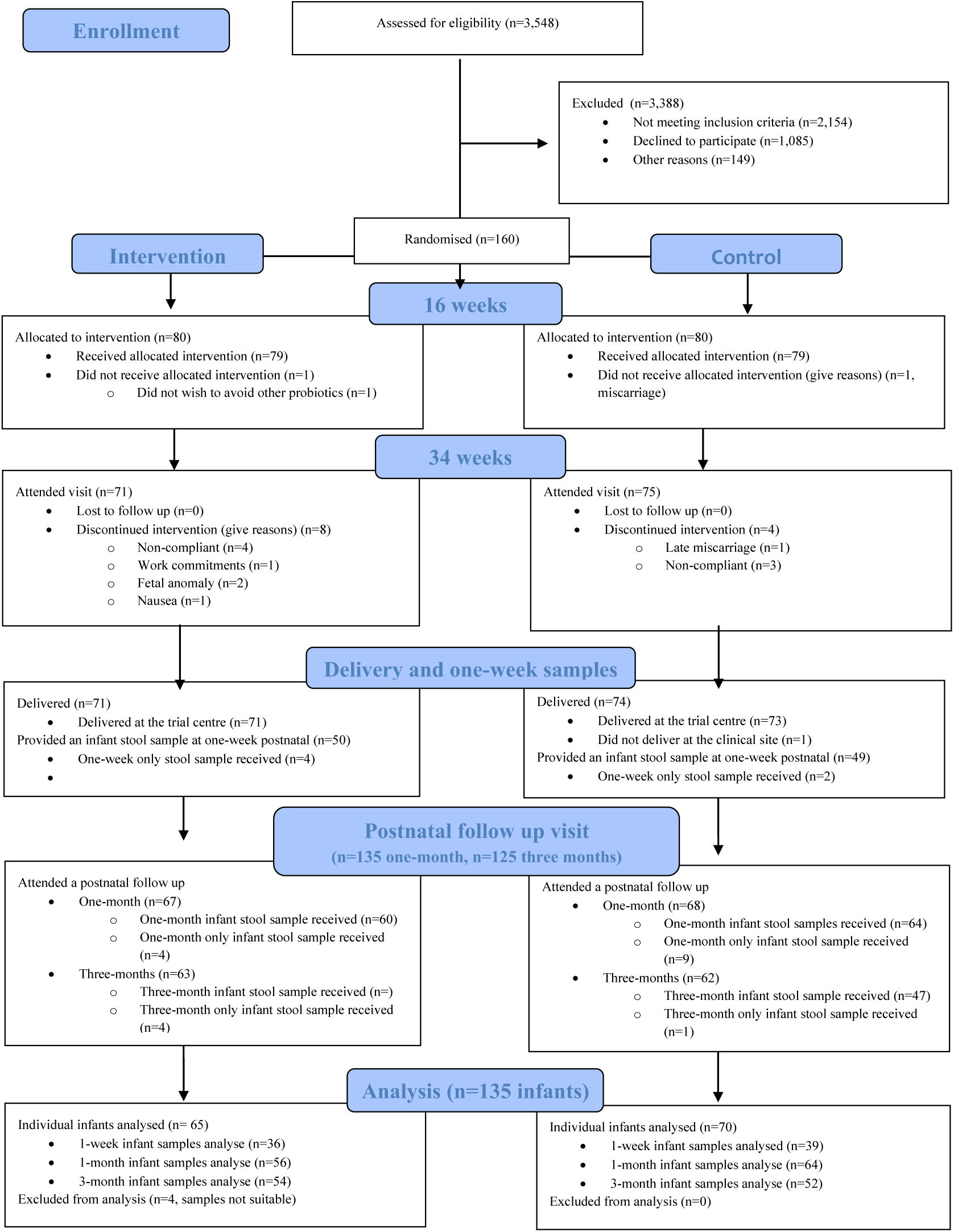
Participant recruitment and intervention flowchart. CONSORT Flow diagram the MicrobeMom study. There were 135 infants (65 in the intervention and 70 in the control group) for whom we received a stool sample for analysis of the primary outcome at either the 1-week, 1-month, or 3-month timepoint. Of these, 24 infants had one sample (six at 1-week, 13 at 1-month and five at 3-months). An additional 58 infants had two samples (12 with both 1-week and 1-month samples, and 42 with both 1-month and 3-month samples). Finally, a further 53 infants had stool samples at all three timepoints. In total, 300 infant stool samples were analysed (74 at 1-week, 120 at 1-month and 106 at 3-months).

### Intervention

At the randomisation visit (approximately 16 weeks’ gestation) participants were allocated to either the intervention or control group. Block randomisation was determined by an independent statistician. Randomly permuted block sizes were generated, and study group allocation was placed in an envelope with the corresponding unique participant identifier. The intervention group received a daily oral supplement of *B. breve* 702258 (all containing a minimum of 1×10^9^ colony forming units; designated from here as “probiotic”) until three months postpartum, while the second group received an otherwise identical placebo (Fig. 1). The supplements were provided by the PrecisionBiotics Ltd. Researchers and participants remained blinded to study allocations until all results were analysed.

### Data collection and trial management

Demographic information collected at the baseline visit included maternal age, ethnicity, parity, and current smoking status via self-report and electronic medical records. Highest level of educational attainment was assessed through a questionnaire. Economic advantage was assessed using the Pobal Haase-Pratschke (HP Pobal) Deprivation Index^29, 30^. Participants were classified as economically advantaged based on a score greater than zero. Baseline weight was measured to the nearest 0.1kg with the mother in light clothing, using a SECA weighing scales (SECA gmbh & co. kg. Hamburg, Germany). Height was measured to the nearest 0.1 cm, using a wall mounted stadiometer, after removal of footwear. These values were used to calculate maternal body mass index (BMI). The trial steering committee met monthly and reviewed study activities.

### Biological samples

#### Maternal

Maternal venous blood samples were collected at 16-weeks’ and 34-weeks’ gestation after an overnight fast. Maternal breastmilk samples were collected at 1-month postpartum. Maternal stool samples were collected at 16 weeks’ and at 34-weeks’ gestation. Participants were asked to provide a whole bowel motion produced within 24 hours before their visit that was stored in a cool dry place until their appointment. A sub-sample of stool (1 g) was taken and added to 7 ml RNAlater. As an environmental control, RNAlater was added to stool-free sample pots and frozen. The remaining sample was stored in containers at −80°C.

#### Infant

Neonatal stool samples at 1-week, 1-month, and 3-months postpartum were collected from infant nappies. A sub-sample of stool (1 g) was taken and added to 7 ml RNAlater. As an environmental control, RNAlater was added to stool-free sample pots and frozen. The remaining sample was stored in containers at -80°C. PCR analysis was carried out samples to 3-months of age, culture work was performed on limited samples to 3-months of age, and metagenomic sequencing was performed up to 1-month of age.

### Primary outcome

The primary outcome was transmission of the bifidobacterial strain to infant stool. Confirmation of transfer was considered when the results of at least two methods of analysis (detailed below) indicated a positive result.

### Nucleic acid extraction

DNA was extracted from all RNAlater fixed stool samples within 8 weeks of sample collection using the AllPrep DNA/RNA kit (Qiagen). Briefly, 100 mg of faecal sample was centrifuged at maximum speed for 10 min and excess RNAlater and supernatant was removed. The pellet was resuspended with 100 μl bacterial lysis buffer (30 mM Tris-HCl, pH 8.0, 1mM EDTA plus 15 mg ml-1 lysozyme) and 10 μl proteinase K (20 mg ml-1)). Following incubation, 1.2 ml Qiagen RLT Plus buffer containing 1% β-mercaptoethanol was added to the sample and vortexed briefly. Samples were transferred to 2 ml sterile bead beating tubes filled with 1 ml of 0.1 mm glass beads and subjected to bead beating for 3 min. 700 μl of lysate was added to a QIAshredder spin column (Qiagen) and centrifuged at maximum speed for 2 min. DNA was extracted from the lysed sample using the AllPrep kit (Qiagen) as per manufacturer’s instructions. Negative control extraction blanks were included for every 50 samples. DNA was extracted from breast milk samples as previously described^31^.

### Metagenomic sequencing and quality control

DNA was quantified and normalized to a concentration of 0.2 ng/ml using the Qubit™ dsDNA HS Assay Kit. Sequencing libraries were prepared following the Nextera XT DNA Library Preparation Kit (Illumina) protocol and then pooled to a concentration of 2 mM. The libraries were sequenced using an Illumina NextSeq platform with a 2×300 bp paired-end kit, generating raw data sets that were subjected to base calling with Illumina’s bcl2fastq software (v 2.19). Adapter removal and quality trimming were performed using TrimGalore (v 0.6.0), which utilized Cutadapt 42 (v 2.6) and FastQC (v 0.11.8) with default parameters. The resulting reads were aligned to the human genome with Bowtie2 43 (v 2.3.4), followed by the removal of aligned reads with samtools 44 (v 1.9) and conversion from BAM to fastq format with bedtools 45 (v 2.27.1). Interleaved fastq files were generated using BBMap (v 38.22). Samples without taxonomic assignment were removed, and samples with fewer than 100,000 post-filtering reads were excluded as they did not meet the maximum number of reads observed in negative control samples. Lastly, singleton samples (maternal samples without corresponding infant samples) were removed.

### Bioinformatic analysis

Compositional profiling was carried out with HUMAnN3 (v3.0) using MetaPhlAn3 database version v30. Metagenomes were assembled with metaSPAdes (v. 3.14), and assembly statistics were calculated using the assembly_stats python library (v. 0.1.4). To recover metagenome-assembled genomes (MAGs), paired fastq reads were aligned to assembled metagenomes by Bowtie2 with default parameters. The resulting SAM files were converted to BAM format and sorted using Samtools, and contig depth was calculated using the jgi_summarise_bam_contig_depths script bundled with MetaBat2 (v. 2.12.1). MetaBat2 was also used for contig binning and the lineage_wf workflow of CheckM (v. 1.0.18) was used to evaluate the quality of metagenomic bins. Only those bins with completeness ≥ 90% and contamination < 5% were considered “high quality” and retained for downstream analyses.

Community-level microbiome analysis was performed using the vegan package in R with the MetaPhlAn3 species table as input. Alpha (within-sample) diversity was calculated using Shannon, Simpson, and observed species indexes. Statistical difference in diversity between trial groups was tested with multiple comparison using Bonferroni. Beta (between-sample) diversity was calculated using Bray-Curtis dissimilarity with significant differences between trial groups tested with PERMANOVA using vegan’s adonis function.

### Selective isolation of *Bifidobacterium* spp

Stool aliquots were retrieved from storage and allowed to thaw at room temperature before resuspension in phosphate buffered saline (PBS; Sigma Aldrich) + 0.06% cysteine-HCL (Sigma Aldrich) at a concentration of 0.1 mg ml^-1^. Samples were thoroughly resuspended by vortexing and then serial ten-fold dilutions performed and plated onto modified MRS (Difco™ Lactobacilli MRS Broth, BD) broth containing either lactose (Merck), 2’FL, or LNnT (both of these human milk oligosaccharides were obtained through the donation programme of Glycom A/S, a DSM group company) as sole sugar. Breast milk aliquots were allowed to thaw on ice for 30 min prior inoculation into MRS, supplemented with lactose and 0.06% cysteine-HCl. After 24 hours incubation in anaerobic conditions at 37 °C, the cell suspension was plated in modified MRS broth containing lactose. Individual colonies were screened in both cases and isolated strains were confirmed to belong to the *Bifidobacterium* spp. by a biochemical assay to detect fructose-6 phosphate phosphoketolase activity as previously described^32^.

### Bacterial genomic DNA extraction

Overnight bacterial cultures of a single colonie were grown in mMRS + 0.5% (w/v) lactose and DNA extracted using GenElute bacterial DNA isolation kit (Merck) according to manufacturer’s instructions. Briefly, cells were lysed using Gram positive lysis solution supplemented with 2.115 × 10^6^ units ml^-1^ lysozyme from chicken egg white (Merck) and incubated for 30 min at 37℃. RNA was degraded using RNase A provided in the kit. Posterior incubation in lysis solution C plus proteinase K (20 mg ml^-1^) was performed followed by the addition of 96% (v/v) ethanol. The lysate was then transferred to a GenElute Nucleic Acid Binding Columns. Following binding and washing, DNA was eluted in 110 µl of Elution solution (10 mM Tris-HCl, 0.5 mM EDTA, pH 9.0).

### Genomic DNA sequencing, assembly, and annotation

The Illumina HiSeq platform was used to sequence gDNA (GenProbio s.r.l., Parma, Italy). Raw sequence reads were assembled into contigs using Spades (v 3.1.1). Prodigal (v 2.6.3) was used to predict open reading frames (ORF) that were then aligned to a combined bifidobacterial genome-based database with a BLASTP v2.6.0 (cut-off E-value of 0.0001) and a MySQL relational database used to assign annotations. Transfer RNA genes were identified employing tRNAscan-SE v1.3.1 and ribosomal RNA genes were detected based on the software package Rnammer v1.2 supported by BLASTN v2.6.0.

### Strain classification and SNP region phylogenetic reconstructions

Taxonomical classification of strains was based on whole genome comparison to a local database of genomes of 66 *Bifidobacterium* species using fastANI v1.3. A threshold of 98% average nucleotide identity was used to classify strains. A Phylogenetic tree based on core single nucleotide polymorphisms was SNP generated as follows. Raw paired end reads were trimmed with trimmomatic (v. 0.36) with the following options “-phred33 ILLUMINACLIP:TruSeq3-PE.fa:2:30:10 LEADING:3 TRAILING:3 SLIDINGWINDOW:5:20 MINLEN:50”. Snippy (v. 4.4.5) was used to predict SNPs for each strain compared to a species-specific reference genome and to generate an alignment of core SNPs. Gubbins (v. 3.0.0) was used to identify and remove putative areas of recombination with the following options “-m 4 -b 4000 --first-tree-builder fasttree”. RAxML was used to generate a phylogenetic tree with RAxML as follows: 20 maximum likelihood (ML) trees (parameters: -m ASC_GTRGAMMA --asc-corr=lewis -p 12345 -N 20) and 100 bootstrap trees (parameters: -m ASC_GTRGAMMA --asc-corr=lewis -p 12345 -x 12345 -N 100) were used to generate the best tree (parameters: -m ASC_GTRGAMMA --asc-corr=lewis -p 12345 -f b). Phylogenetic trees were visualised and annotated in iTOL (https://itol.embl.de/). All genomes of isolated *Bifidobacterium breve* strains were compared to the genome of *B. breve* 702258 with fastANI. Strains that had an average nucleotide identity (ANI) >= 99.9 % when compared to the *B. breve* 702258 genome were deemed to be the same strain Finally, the phylogenetic tree of the core single nucleotide polymorphism (SNP) regions of was examined to identify if these strains formed monophyletic group with *B. breve* 702258.

### PCR detection of *B. breve* 702258

To detect the presence of *B. breve* 702258 in samples, a primer set specific to a distinct CRISPR spacer sequence within the 702258 genome was designed (qBRV-F | GGCGAACAATTTCAATC; qBRV-R | GGTTGAGTATCGTATTGG), along with a product specific probe (250 nM PrimeTime 5’ 6-FAM/ZEN/3’IB FQ CTCCGAACCATTTCAATCCAC; IDTDNA). Primer efficiency was determined to be >95% with an R^2^ of 0.9996. Briefly, DNA was normalized to 0.2 ng µl^-1^ and 3 µl used as template with 0.8 µl of each primer (10 µM), 0.4 µl probe (10 µM), 10 µl Luna Universal probe qPCR master mix (NEB), and 5 µl water. Reactions were carried out in 96-well plates using Lightcycler 480 (Roche) with initial denaturation for 60 s at 95°C, followed by 45 cycles of denaturation for 15 s at 95°C and extension for 30 s at 60°C with signal detection using LC480 FAM filter. Positive signal was regarded as ct value less than negative control of sterile water. A positive template DNA control (genomic DNA extracted from *B. breve* 702258) was included for all plate runs. All suspected positive detections of 702258 determined using qPCR were confirmed using regular PCR with product visualization on 1.5% agarose gel. For this PCR, a second set of primers targeting the same CRISPR spacer region were used (ION-F | ATTCGTATACAGGGCGGTTG; ION-R | GCTCGAATGCGGTTGAGTAT). Briefly, 1 µl DNA was mixed with 0.5 µl of each primer, 12.5 µl Accuzyme (Bioline), and 10.5 µl water and cycled with initial denaturation for 2 min at 94°C, and 30 cycles of 1 min at 94°C, 25 s at 58°C, 20 s at 72°C, followed by final elongation for 1 min at 72°C.

### Secondary outcomes

Prespecified secondary outcomes included maternal lipids, glucose, insulin resistance (homeostatic model assessment (HOMA-IR)), and C-reactive protein. Fasting glucose samples were analysed at the clinical site after the shortest possible time interval post sample collection^33, 34^. The Beckman Coulter AU680 analyser and the Hexokinase method was employed for this measurement. The rest of the samples taken were centrifuged at 3000 rpm for 10 min. Aliquots were stored at −80°C. Total cholesterol, high-density lipoprotein (HDL) cholesterol, and triglyceride concentrations were analysed on a Roche c702 analyser (Roche Diagnostics). Low density lipoprotein (LDL) cholesterol was estimated using the Friedewald equation^33^. Serum measurements of insulin were determined Insulin was quantified by automated immunoassay (Roche, E170) with typical coefficients of variance <5%. The HOMA-IR was calculated from fasting glucose and insulin levels using the following equation: (fasting insulin (Mu/l) x fasting glucose (mmol/l))/22·5^34^.

Additional outcomes included maternal and infant microbiome (composition and abundance), inflammatory markers (C3 Complement (C3), Interleukin 6 (IL-6), Tissue-necrosis factor alpha (TNF-Alpha)), lipoproteins (Apo-a, Apo-B100 and Lipoprotein a), Glycosylated Haemoglobin (HbA1c) and pregnancy outcomes. Kits from R&D systems, a bio-techne brand were used to measure TNF-alpha. The electrochemiluminescence immunoassay (ECLIA) was used for the quantitative determination of IL-6 using the Roche cobas e411. Complement C3 was completed using the Abbott Architect c4000. Rate of gestational weight gain per week was calculated and compared to the institute of medicine guidelines^35^.

Participants who had an oral glucose tolerance test (OGTT in late pregnancy (28-30 weeks) (n=22, 16·3%) were assessed against the International Association of Diabetes and Pregnancy Study Group diagnostic criteria^36^. Pregnancy-induced hypertension, pre-eclampsia, mode of delivery, preterm birth (delivery of a liveborn infant <37-weeks’ gestation), post-partum haemorrhage (> 500ml blood loss), placental weight, birthweight (including low birth weight (<2500 g) and macrosomia (>4000 g)) and neonatal intensive care unit admission were recorded from medical records. Customised birthweight centiles were calculated using the Gestation Network’s Bulk Calculator (version 8·0·6·1, 2020)^37^. Rates of small for gestational (SGA) (<5^th^ or 10^th^ centile) and large for gestational age (LGA) (>90^th^ centile) were determined. Breastfeeding practices and antibiotic use were further assessed at postnatal visits.

### Adherence

Participants were asked if agreed they took the capsules every-day using a five-point Likert scale, at 34-weeks’ gestation, 1-month, and 3-months postpartum, as done previously^38^. Participants were also asked to return any unused capsules at the end of the study and received fortnightly text messages.

### Sample size and power

A power analysis determined 60 participants per group were required to provide at least one stool sample up to 3 months of age to detect a difference in the primary outcome at a significance level of 5% and a target power of 80%. In the absence of available literature, this was based on an expected colonisation rate in the placebo of 0% and 15% in the intervention group.

### Statistical analysis

Continuous variables were assessed for normality and all non-normally distributed data underwent log^10^ or exponential transformation prior to analysis. If normality was not achieved, the appropriate non-parametric test was used. Continuous variables are presented as mean (standard deviation) or median and interquartile range (25th, 75th centile) for non-normally distributed data. Categorical variables are presented as number and frequency (%). Primary and secondary outcomes were compared between groups. Comparison statistics were generated through independent sample t-tests using parametric or normally distributed log-transformed data. Mann-Whitney U test was used for non-parametric data. Chi-square (*χ*2) tests were used for categorical variables, and Fishers’ exact tests were used when assumptions for chi-square were violated. In the case of significant findings, relevant baseline information between study groups were compared. Results for the primary outcome include relative risk (RR) and 95% Confidence Intervals (95% CI). Within group comparisons were generated through paired-sample t-tests or Wilcox tests. Absolute mean differences are reported. This study employed an intention to treat analysis. Tests were performed using IBM Statistical Package for Social Sciences software for Windows, version 26·0 (SPSS Inc, Chicago IL) for all except the RR values, which were generated using Rstudio version 4·1·3 (2022-03-10), package epiR.

## Results

The consort flow diagram is shown in Fig. 1. There were 135 infants (65 in the intervention and 70 in the control group) for whom we received a stool sample at either the 1-week, 1-month, or 3-month timepoint. Of these, 24 infants had one sample (6 at 1-week, 13 at 1-month and 5 at 3-months), 58 had two samples (12 had 1-week and 1-month samples and 42 had 1-month and 3-month samples) and 53 infants had samples at all three timepoints. In total, 300 infant stool samples were analysed (74 at 1-week, 120 at 1-month and 106 at 3-months). Demography of study participants is shown on Table 1. At discharge from hospital, 83·2% (n=110) of infants were receiving some breastmilk. In post-hoc analysis, the probiotic group were younger (mean (SD) age of 32·66 (3·89)) than placebo (mean (SD) age 34.36 (2·78), *p*=0·011) and had less multiparity (n=21 (32·3%) vs n=37 (57·0%), *p* = 0·012).

**Table 1:**
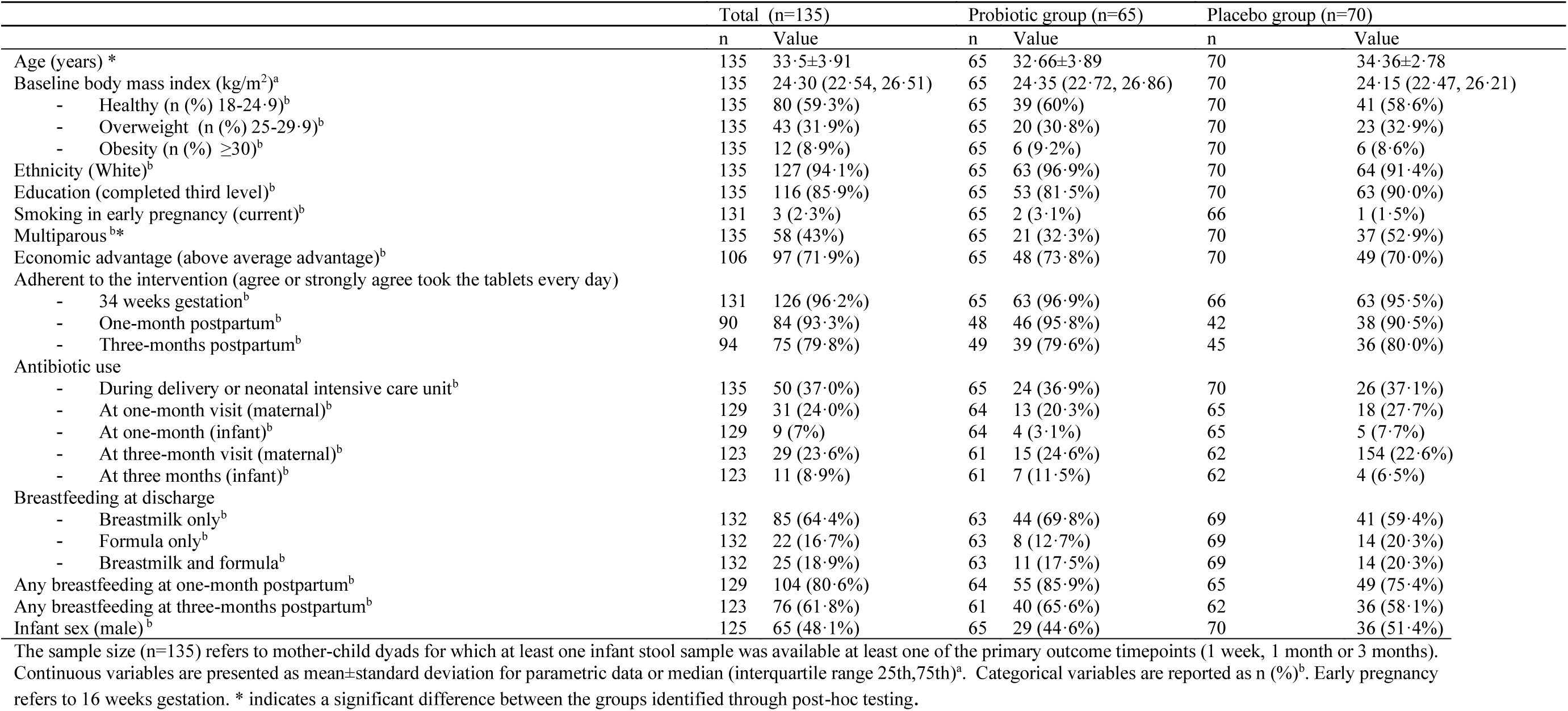
Maternal and infant characteristics in the MicrobeMom Randomised Controlled Trial

|  | Total (n=135) |  | Probiotic group (n=65) |  | Placebo group (n=70) |  |
| --- | --- | --- | --- | --- | --- | --- |
|  | n | Value | n | Value | n | Value |
| Age (years) * | 135 | 33.5±3.91 | 65 | 32.66±3.89 | 70 | 34.36±2.78 |
| Baseline body mass index (kg/m<sup>2</sup>)<sup>a</sup> | 135 | 24.30 (22.54, 26.51) | 65 | 24.35 (22.72, 26.86) | 70 | 24.15 (22.47, 26.21) |
| - Healthy (n (%) 18-24.9)<sup>b</sup> | 135 | 80 (59.3%) | 65 | 39 (60%) | 70 | 41 (58.6%) |
| - Overweight (n (%) 25-29.9)<sup>b</sup> | 135 | 43 (31.9%) | 65 | 20 (30.8%) | 70 | 23 (32.9%) |
| - Obesity (n (%) ≥30)<sup>b</sup> | 135 | 12 (8.9%) | 65 | 6 (9.2%) | 70 | 6 (8.6%) |
| Ethnicity (White)<sup>b</sup> | 135 | 127 (94.1%) | 65 | 63 (96.9%) | 70 | 64 (91.4%) |
| Education (completed third level)<sup>b</sup> | 135 | 116 (85.9%) | 65 | 53 (81.5%) | 70 | 63 (90.0%) |
| Smoking in early pregnancy (current)<sup>b</sup> | 131 | 3 (2.3%) | 65 | 2 (3.1%) | 66 | 1 (1.5%) |
| Multiparous<sup>b*</sup> | 135 | 58 (43%) | 65 | 21 (32.3%) | 70 | 37 (52.9%) |
| Economic advantage (above average advantage)<sup>b</sup> | 106 | 97 (71.9%) | 65 | 48 (73.8%) | 70 | 49 (70.0%) |
| Adherent to the intervention (agree or strongly agree took the tablets every day) |  |  |  |  |  |  |
| - 34 weeks gestation<sup>b</sup> | 131 | 126 (96.2%) | 65 | 63 (96.9%) | 66 | 63 (95.5%) |
| - One-month postpartum<sup>b</sup> | 90 | 84 (93.3%) | 48 | 46 (95.8%) | 42 | 38 (90.5%) |
| - Three-months postpartum<sup>b</sup> | 94 | 75 (79.8%) | 49 | 39 (79.6%) | 45 | 36 (80.0%) |
| Antibiotic use |  |  |  |  |  |  |
| - During delivery or neonatal intensive care unit<sup>b</sup> | 135 | 50 (37.0%) | 65 | 24 (36.9%) | 70 | 26 (37.1%) |
| - At one-month visit (maternal)<sup>b</sup> | 129 | 31 (24.0%) | 64 | 13 (20.3%) | 65 | 18 (27.7%) |
| - At one-month (infant)<sup>b</sup> | 129 | 9 (7%) | 64 | 4 (3.1%) | 65 | 5 (7.7%) |
| - At three-month visit (maternal)<sup>b</sup> | 123 | 29 (23.6%) | 61 | 15 (24.6%) | 62 | 154 (22.6%) |
| - At three months (infant)<sup>b</sup> | 123 | 11 (8.9%) | 61 | 7 (11.5%) | 62 | 4 (6.5%) |
| Breastfeeding at discharge |  |  |  |  |  |  |
| - Breastmilk only<sup>b</sup> | 132 | 85 (64.4%) | 63 | 44 (69.8%) | 69 | 41 (59.4%) |
| - Formula only<sup>b</sup> | 132 | 22 (16.7%) | 63 | 8 (12.7%) | 69 | 14 (20.3%) |
| - Breastmilk and formula<sup>b</sup> | 132 | 25 (18.9%) | 63 | 11 (17.5%) | 69 | 14 (20.3%) |
| Any breastfeeding at one-month postpartum<sup>b</sup> | 129 | 104 (80.6%) | 64 | 55 (85.9%) | 65 | 49 (75.4%) |
| Any breastfeeding at three-months postpartum<sup>b</sup> | 123 | 76 (61.8%) | 61 | 40 (65.6%) | 62 | 36 (58.1%) |
| Infant sex (male)<sup>b</sup> | 125 | 65 (48.1%) | 65 | 29 (44.6%) | 70 | 36 (51.4%) |

The sample size (n=135) refers to mother-child dyads for which at least one infant stool sample was available at least one of the primary outcome timepoints (1 week, 1 month or 3 months).
Continuous variables are presented as mean±standard deviation for parametric data or median (interquartile range 25th,75th)<sup>a</sup>. Categorical variables are reported as n (%).<sup>b</sup> Early pregnancy refers to 16 weeks gestation. \* indicates a significant difference between the groups identified through post-hoc testing.

### Primary Outcome

The primary outcome of detection of the supplemented strain was confidently identified by at least two methods in two different infants within the probiotic group and none in the placebo group (Table 2). These two mothers both had a previous liveborn infant, were White Irish and had a BMI in the overweight category (BMI between 25-29.9kg/m^2^) and strongly agreed they took the probiotic capsules every day in late pregnancy. Both infants were male and born at term, one by planned prelabour caesarean delivery and one by spontaneous vaginal delivery. Both were breastfed on hospital discharge.

**Table 2:**
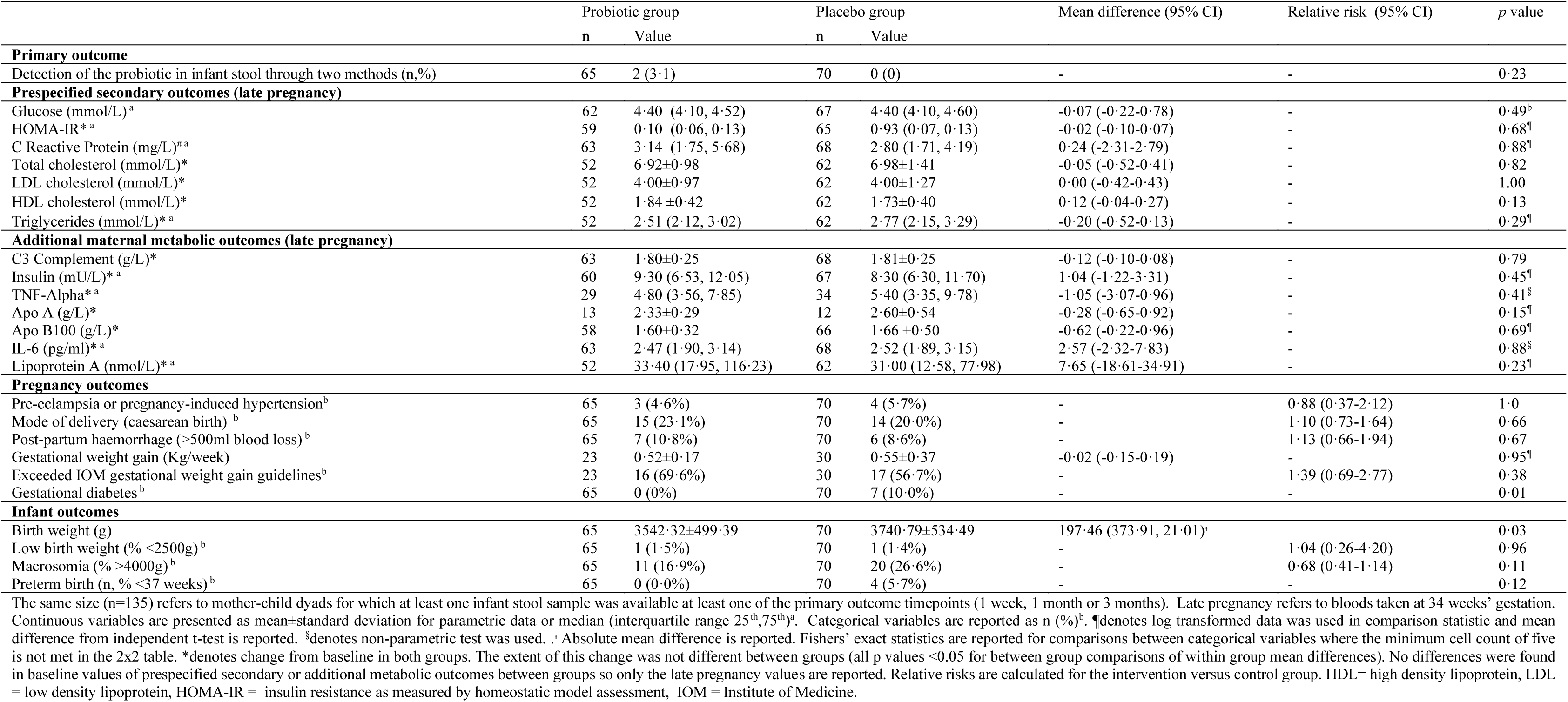
Primary and secondary outcomes after the intervention in the MicrobeMom Study

### Detection of the probiotic strain

Using PCR at any time-point suggestive evidence of the *B. breve* 702258 strain was found in five infants, three in the probiotic (4·6%) and two in the placebo group (2·9%) (Fig. 2 a). Of these, one infant in the probiotic group had a positive PCR at all three timepoints. Another infant from the probiotic group had two positive samples but lacked a 1-week sample. The third infant in the probiotic group had a positive PCR detection at the 3-month timepoint but not at 1-month and did not have a 1-week sample. The two infants in the placebo group had samples tested at all three time-points, but for one, the positive signal was found in the 3-month sample only and for the other, at 1-month only. Using a targeted culture-based approach, 60 distinct *B. breve* isolates were obtained from infant stool, however genome sequencing revealed only two of these were highly similar (>99·99% identity) to the supplemented strain (Fig 2 c). Both were from infants in the probiotic group (detection rate in probiotic group of 3·1%), at either 1-month or 3-months. Both samples also had a respective positive PCR result. Culture analysis did not find any positive isolates in the placebo group (0%). There were 37 distinct occurrences of *B. breve* in infant stool detected. Only one metagenomic assembled genome (MAG) from a single stool sample in the probiotic group was identified with a high similarity to the genome of the supplemented strain (Fig. 2 b). This was in a 1-month infant sample that also had both a positive PCR detection and to which a confirmed isolate had been recovered and sequenced at the 3-month timepoint.

**Figure 2:**
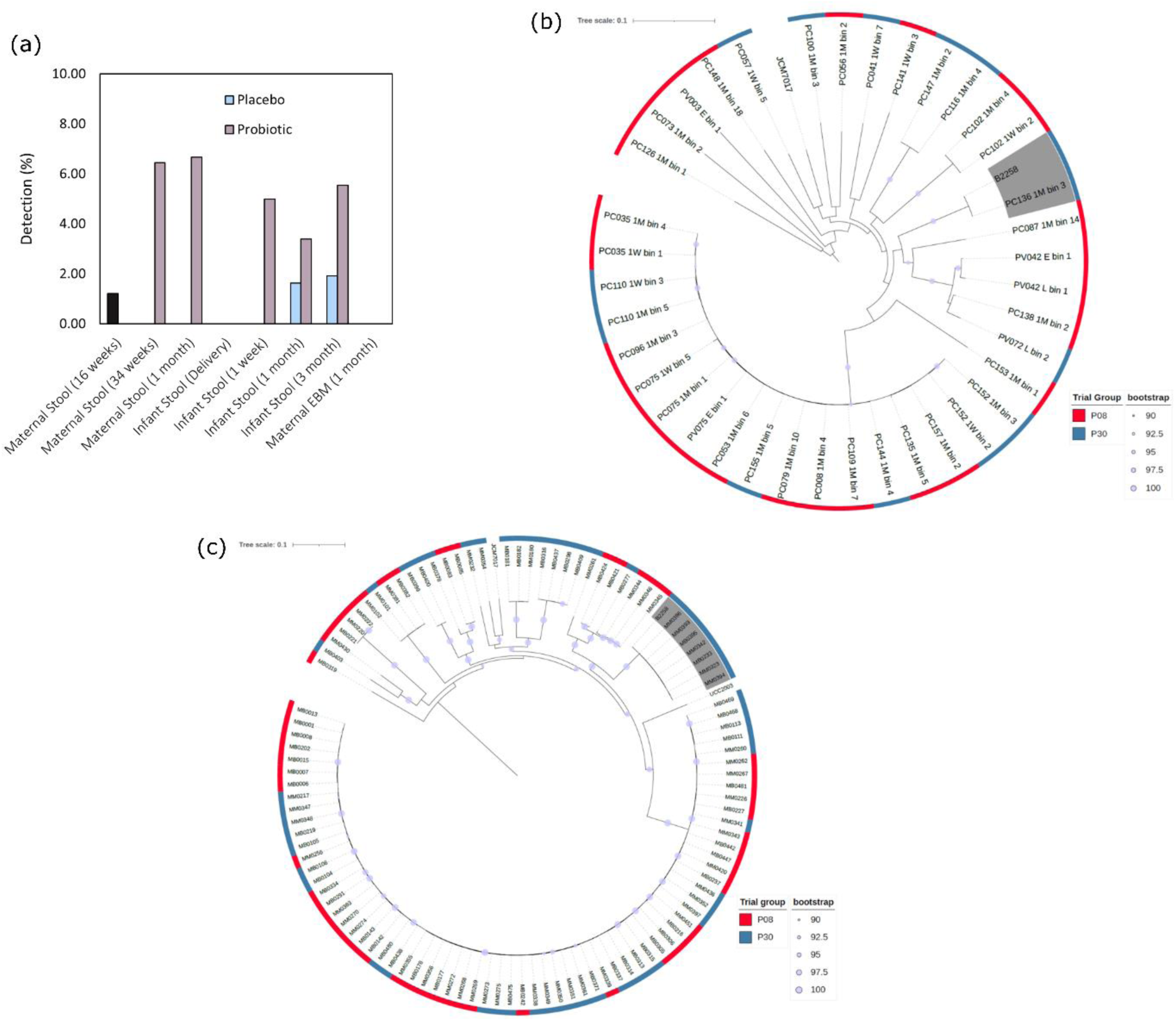
Detection of supplemented *B. breve* 702258. PCR analysis to detect the strain-specific marker gene of the probiotic strain. Bar charts illustrate the percentage of samples in each trial group that had a positive detection of B. breve 702258 for each sample type. (b) Maximum likelihood phylogenetic tree showing the relatedness of metagenome-assembled genomes (MAGs) recovered from the infant stool to the genome of B. breve 702258. (c) Phylogenetic tree showing the genomic relatedness of cultured B. breve isolates from infant stool to the B. breve 702258 genome. Phylogenetic trees were based on the alignment of core SNP regions of all genomes shown.

### Secondary Outcomes

No differences were found in baseline values of prespecified secondary or additional metabolic outcomes between groups so only the late pregnancy values are reported. No differences were noted in metabolic factors in late pregnancy (Table 2). All women with GDM were in the placebo group (RR 0·16, 95% CI 0·01, 2·47, *p*=0·121) and there was a difference in HbA1c at baseline between intervention and control (29·88 (2·44) vs 31·09 (2·41), *p*=0·006). Both groups experienced an increase in their HbA1c with gestation, but the extent of this change was not different between groups (*p*=0·888) (supplementary Table 1). There was also higher incidence of large for gestational age infants in the placebo group (n=19, 14·7%) compared to the probiotic group (n=6, 9·4%, RR 0·32, 95% CI 0·14, 0·75, *p*=0·004).

Using PCR analysis, the probiotic strain was detected in nine maternal samples, one at randomisation, four at 34-weeks’ gestation, and four at 1-month postpartum (Fig. 2 a). Only two of these (both at 34-weeks and in the intervention group) had cultured bacterial isolates from the same sample with an ANI score of >99.99% to *B. breve* 702258. The maternal sample with a positive PCR at randomisation did not show evidence of the probiotic by either culture work or metagenomics. A total of 55 *B. breve* isolates were cultured from maternal stool and breastmilk samples. Of these, four isolates had a genome identity of >99·99% to the probiotic. Only one of these samples came from a mother that did not have a positive PCR. A single isolate was recovered from breastmilk at 1-month that matched the supplemented strain. This was from a mother/infant dyad which had a positive detection in the infant stool at the same timepoint. This infant was fed some breastmilk (and formula) from delivery but had moved to formula only by three months. No *B. breve* MAG with a similarity to the supplemented strain were recovered from maternal stool metagenomic sequencing. Taken together, only two mothers from the probiotic group (2/65, 3·1%) and none in the control group had evidence of the probiotic strain confirmed in their stool through two methods *p=*0·230. Whilst PCR analysis determined the presence of *B*. *breve* 702258 in two different mother/infant dyads, this was not reproduced by the other methods at the same timepoint. Independent of time point and sample site however, there was evidence for the strain in a single dyad by both PCR and culture.

There was no difference in microbial community diversity in both maternal and infant samples (Fig. 3 a & b). Supplementation also did not influence the community structure as measured by Bray-Curtis (Fig. 3 c). In the maternal samples, the overall relative abundance of *Bifidobacterium* (including *B. breve*) remained stable across all timepoints and there was no difference between groups (Fig. 3 d). The relative abundance of total *Bifidobacterium* in infant samples increased over time but this was not influenced by the probiotic (Fig. 4 d & e).

**Figure 3:**
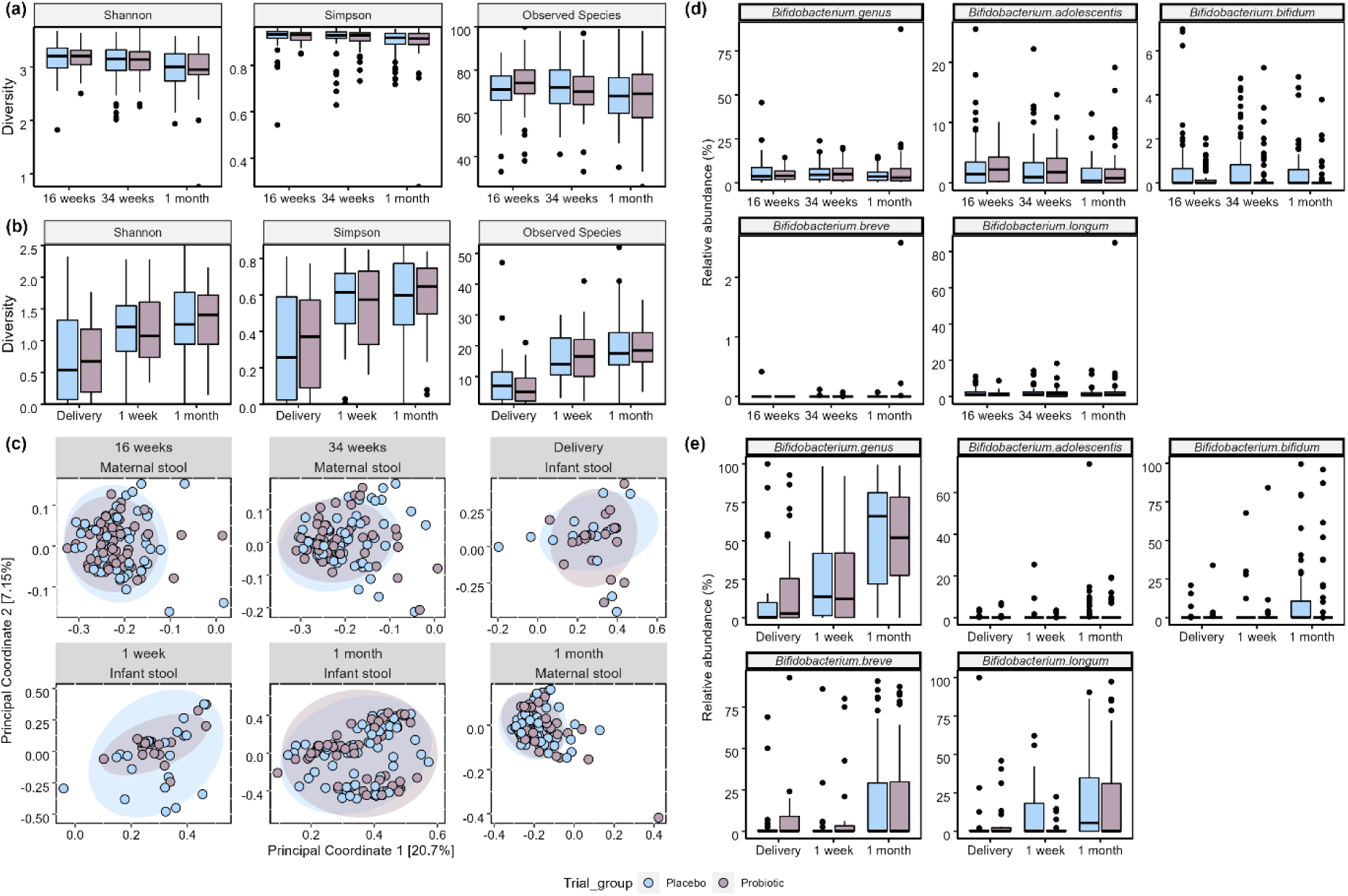
Microbiome composition analysis of the maternal & infant stool microbiome Alpha diversity of the maternal stool (a) & infant stool (b) microbiome as determined by Simpson, Shannon, and Observed species. Beta diversity of both the maternal & infant stool microbiome (c) as determined using Bray-Curtis dissimilarity measure. The relative abundance of overall Bifidobacterium and the top 4 individual Bifidobacterium species in the maternal (d) and infant (e) stool compared between trial groups.

### Comment

#### Principal findings

Our data suggests mother to child transmission in two cases. There were no differences in pre-specified secondary outcomes. The placebo group had higher incidence of GDM and more LGA infants, but this may be explained by higher age, multiparity and baseline HbA1c in the control group.

### Results in context of what is known

We hypothesised that maternal probiotic supplementation would support transfer of the probiotic to the infant. There is some other albeit limited evidence to suggest that this is possible, and the mechanisms are yet to be determined^23, 39^. In our study, there was a confirmed culture of *B.* breve 702258 from a maternal breastmilk sample for which there was a detection and isolation of the strain from the respective infant. Whilst tempting to consider this as a mode of transmission, the rare occurrence limits any certainty. The overall detection rate of transmission was lower than expected.

In their study of mothers of infants at high risk of eczema, Lahtinen *et al* were the first to show that an antenatal probiotic (*Lactobacillus rhamnosus* GG) given to mothers from 36-weeks’ gestation can alter the bifidobacterial composition of infants^39^. Although they did not find transfer of the probiotic from mothers to children, they found infants born to mothers in the intervention group had higher colonisation with *B. longum*. This suggests a potential favourable effect^40^. In contrast, we did not see an impact on infant, or maternal, microbial composition, although our study was not powered to any subtle effects.

We provided a single probiotic strain whilst some others have supplemented with multiple strains^22, 41^. In a study of antenatal probiotic milk (containing *L. rhamnosus GG*, *Lactobacillus acidophilus La-5*, and *Bifidobacterium animalis subsp. lactis Bb-12*), Dotterud *et al.* found that only *L. rhamnosus* GG was detected in infant faecal samples at ten-days and three-months of age^42^. This suggests effects on the maternal and infant microbiome may be both strain-specific and time-dependent. The selection of an alternative strain that has a high transfer efficiency and can establish in the maternal microbiome will likely provide greater transfer from mothers to infants.

We found that maternal intake of *B. breve* 702258 did not alter maternal biomarkers. The dose of the probiotic (10^9^ CFU) likely does not explain the lack of findings as a dose of at least 10^7^ CFU is considered large enough to confer a benefit and doses from 10^5^ – 10^14^ CFU have been used in pregnancy^43^. Although there are plausible mechanistic pathways, such as production of short-chain fatty acids, several systematic reviews suggest no conclusive benefit of probiotic intake on metabolic and inflammatory markers in healthy populations, including preconception and pregnancy^44–46^. A recent meta-analysis of 16 studies of pregnancy probiotic supplementation in relation to GDM specifically however, found positive effects on metabolic status including insulin, fasting glucose, and HOMA-IR^43^. One included study in this review found attenuated increases in LDL cholesterol in women with a new diagnosis of GDM or impaired glucose tolerance with daily probiotic supplementation (*Lactobacillus salivarius* UCC118) until delivery^47^.

In line with previous research, we did not find any effect on mode of delivery or preterm birth^48^. This finding further suggests the safety of probiotic supplementation in pregnancy, although we were not powered to see differences in this outcome. Previous research highlights the potential impact of probiotics on the neonatal gut is influenced by a myriad of factors such as maternal gut, mode of delivery, infant feeding, and commensal bacteria^49, 50^. Our study was an intention to treat analysis therefore all infants were included with data on the primary outcome, regardless of these factors. Future work could explore the impact of probiotic supplementation on transmission in select groups of mother-child dyads.

### Research implications

A major strength of our study is the duration of supplementation^23, 51^.This provides crucial insight into the potential safety of longer-term supplementation during pregnancy. Another strength is the use of multiple robust methods to detect the strain, particularly the use of shotgun sequencing, which is lacking in the previous literature^23, 50, 52^. Since positive detections were obtained during PCR for both randomisation stage and placebo group samples, the specificity of PCR could not be relied upon. Incorporation of both culture work and metagenomics enabled a robust analysis, and subsequently only samples with a positive detection by at least two methods were considered when confirming detection. Whilst metagenomic sequencing can be limited by sequencing read depth, and may not detect bacteria present in low abundance, the use of culture and PCR overcomes this. Future work could continue to employ multiple methods of analysis with even greater read depth.

### Clinical implications

Whilst we detected the probiotic strain in infant stool, the low rates suggest that maternal probiotic supplementation with this strain may not be a scalable intervention to influence the infant gut. The mechanisms of colonisation of the infant gut through maternal probiotic supplementation remain unclear and requires more research. Most previous studies have included the antenatal period only, unlike our study where we supplemented up to 3 months postpartum^23, 41^. Although our study was not powered to identify adverse effects and our population was relatively homogenous, clinicians should note the suggested safety of maternal probiotic supplementation from early pregnancy. Given the potential to positively influence the health of infants throughout the life-course, similar trials with alternate or multi-strain probiotics, or synbiotics, are warranted. The role of other factors such as parity, diet and lifestyle should be explored^53, 54^.

## Conclusion

Direct strain transfer from mothers to infants is possible, though at a low level with this strain. This highlights potential for maternal probiotic supplementation as an intervention to introduce beneficial microbial strains into the infant gut microbiome.

## Supporting information

The study is reported according to the EQUATOR network using the CONSORT guidelines (supplemental file 1)

## Data Availability

. All raw sequencing data used in this study is available in the ENA repository under accession number PRJEB48251 (https://www.ebi.ac.uk/ena/browser/view/PRJEB48251). Additional metadata and species tables are available via GitHub (https://github.com/SligoMicrobe/MicrobeMom).

## Acknowledgements

This publication has emanated from research supported in part by a peer reviewed research grant from Science Foundation Ireland (SFI) under Grant No. 12/RC/2273 and 16/SP/3827 which included a research grant from PrecisionBiotics Group Ltd. The authors would like to acknowledge all participants in the MicrobeMom trial.

